# Leveraging chorionic villus biopsies for the derivation of patient-specific trophoblast stem cells

**DOI:** 10.1101/2022.12.07.22283218

**Authors:** Kaela M. Varberg, Ayelen Moreno-Irusta, Allynson Novoa, Brynne Musser, Joseph M. Varberg, Jeremy P. Goering, Irfan Saadi, Khursheed Iqbal, Hiroaki Okae, Takahiro Arima, John Williams, Margareta D. Pisarska, Michael J. Soares

## Abstract

Human trophoblast stem (**TS**) cells are an informative in vitro model for the generation and testing of biologically meaningful hypotheses. The goal of this project was to derive patient-specific TS cell lines from clinically available chorionic villus sampling biopsies. Cell outgrowths were captured from human chorionic villus tissue specimens cultured in modified human TS cell medium. Cell colonies emerged early during the culture and cell lines were established and passaged for several generations. Karyotypes of the newly established chorionic villus-derived trophoblast stem (**TS^CV^**) cell lines were determined and compared to initial genetic diagnoses from freshly isolated chorionic villi. Phenotypes of TS^CV^ cells in the stem state and following differentiation were compared to cytotrophoblast-derived TS (**TS^CT^**) cells. TS^CV^ and TS^CT^ cells uniformly exhibited similarities in the stem state and following differentiation into syncytiotrophoblast and extravillous trophoblast cells. Chorionic villus tissue specimens provide a valuable source for TS cell derivation. They expand the genetic diversity of available TS cells and are associated with defined clinical outcomes. TS^CV^ cell lines provide a new set of experimental tools for investigating trophoblast cell lineage development.

**Summary statement:** The chorionic villus-derived trophoblast stem cell lines described in this report enhance the genetic diversity of available trophoblast stem cell models and are linked to specific clinical outcomes. They offer a novel set of experimental tools for studying trophoblast cell lineage development and human placentation.

## Introduction

The placenta is a critical organ that allows the fetus to develop within the female reproductive tract (Amoroso, 1968). Specialized functions attributed to the placenta are executed by trophoblast cells (Knöfler et al., 2019, Soares et al., 2018, Turco and Moffett, 2019). The trophoblast cell lineage arises from the initial differentiation event of the embryo (Rossant and Tam, 2017). In the human, trophoblast cells organize into villous and extravillous structures. A villous is comprised of trophoblast and non-trophoblast cell types and includes a self-renewing trophoblast cell population referred to as cytotrophoblast (Aplin and Jones, 2021, Sun et al., 2020). Cytotrophoblast are the progenitors for two differentiated cell populations: syncytiotrophoblast (**STB**) and extravillous trophoblast cells (**EVT**) (Knöfler et al., 2019, Soares et al., 2018, Turco and Moffett, 2019). STB have a fundamental role in regulating nutrient and waste transfer between mother and fetus (Aplin and Jones, 2021), whereas EVT cells exit the placenta and transform the uterus into an environment supporting placental and fetal development (Knöfler et al., 2019, Soares et al., 2018). Failures in placentation are the root cause of an assortment of disorders of pregnancy, including pregnancy loss, preeclampsia, intrauterine growth restriction, and pre-term birth (Brosens et al., 2019, Burton et al., 2016). Regulatory mechanisms underlying human cytotrophoblast self-renewal and differentiation have largely remained elusive.

Recently, conditions for capturing and maintaining human trophoblast stem (**TS**) cells in vitro were described (Okae et al., 2018). Human TS cells have the capacity for self-renewal and differentiation into STB cells or EVT. This in vitro model system has led to the generation of new insights into mechanisms regulating human trophoblast cell development (Bhattacharya et al., 2020, Hornbachner et al., 2021, Ishiuchi et al., 2019, Jaju Bhattad et al., 2020, Muto et al., 2021, Perez-Garcia et al., 2021, Ruane et al., 2022, Saha et al., 2020, Shannon et al., 2022, Sheridan et al., 2021, Takahashi et al., 2019, Varberg et al., 2021). Initial human TS cell lines were derived from blastocysts, or first trimester placental tissue obtained from pregnancy terminations. Establishment of culture conditions for human TS cells led to the derivation of TS cells from pluripotent stem cells (Castel et al., 2020, Cinkornpumin et al., 2020, Dong et al., 2020, Guo et al., 2021, Io et al., 2021, Liu et al., 2020, Wei et al., 2021, Yanagida et al., 2021). These in vitro model systems have provided new insights regarding trophoblast cell development; however, it is unknown whether the origin of these TS cells was compatible with a healthy pregnancy outcome.

Chorionic villus sampling (**CVS**) represents a standard, prenatal care procedure that is performed between 10-14 weeks of gestation (Stranc et al., 1997). Sampling involves the removal of a small amount of chorionic villus tissue, for the purpose of genetic testing. Common indications for retrieving chorionic villus tissue include advanced maternal age, history of infertility, family history (e.g., sibling with genetic anomalies), or an abnormal noninvasive prenatal test result (Pisarska et al., 2016, Stranc et al., 1997). In addition to their use in genetic diagnosis, first trimester chorionic villus tissue has become a robust platform for investigation of placental pathobiology (Flowers et al., 2021, Gonzalez et al., 2021, Pisarska et al., 2016).

In this study, we derived patient-specific human TS cell lines from clinically available chorionic villus tissue. Derivation of TS cell lines from chorionic villus tissue expands the genetic diversity of available human TS cells and importantly is linked to clinical data describing pregnancy outcomes.

## Results

### Derivation of TS cells from chorionic villus biopsies

Chorionic villus biopsies were acquired with patient consent as part of standard medical care. Surplus tissue fragments not used for clinical genetic testing were placed in culture medium used for the expansion of human TS cells (Takahashi et al., 2019); **Fig. 1A**). Tissue pieces attached to type IV collagen-coated tissue culture treated plates (**Fig. 1B**). Cell outgrowths were evident at sites of attachment and expanded over the first several weeks of culture (**Fig. 1C**). Cells and tissue fragments were passaged prior to reaching confluency and replated in 24 well plates. Cell colonies emerged after first passage and steadily expanded with culture medium changes every two days. Colony morphology and growth rates were heterogeneous for the first few passages but became more homogenous after 5-6 passages. The morphology of chorionic villus-derived TS (**TS^CV^**) cells was consistent with the morphology of cytotrophoblast-derived TS cells (**TS^CT^**; **Fig. 1B**, **Fig. 2A**). TS^CV^ cell line expansion was carried out slowly to reduce clonal pressure on derived cells. Cell lines were slowly transitioned into 6 well and 10 cm plate formats after passages 3-4 and 7-8, respectively (**Fig. 1C**). Newly established cell lines were cryopreserved beginning at passage 6. Importantly, TS^CV^ cells tolerated cryopreservation. Revived cells survived, attached, and proliferated for further expansion. Overall, TS^CV^ cell line derivation required approximately three months from sample acquisition to functional assessments of derived lines.

**Fig. 1.**
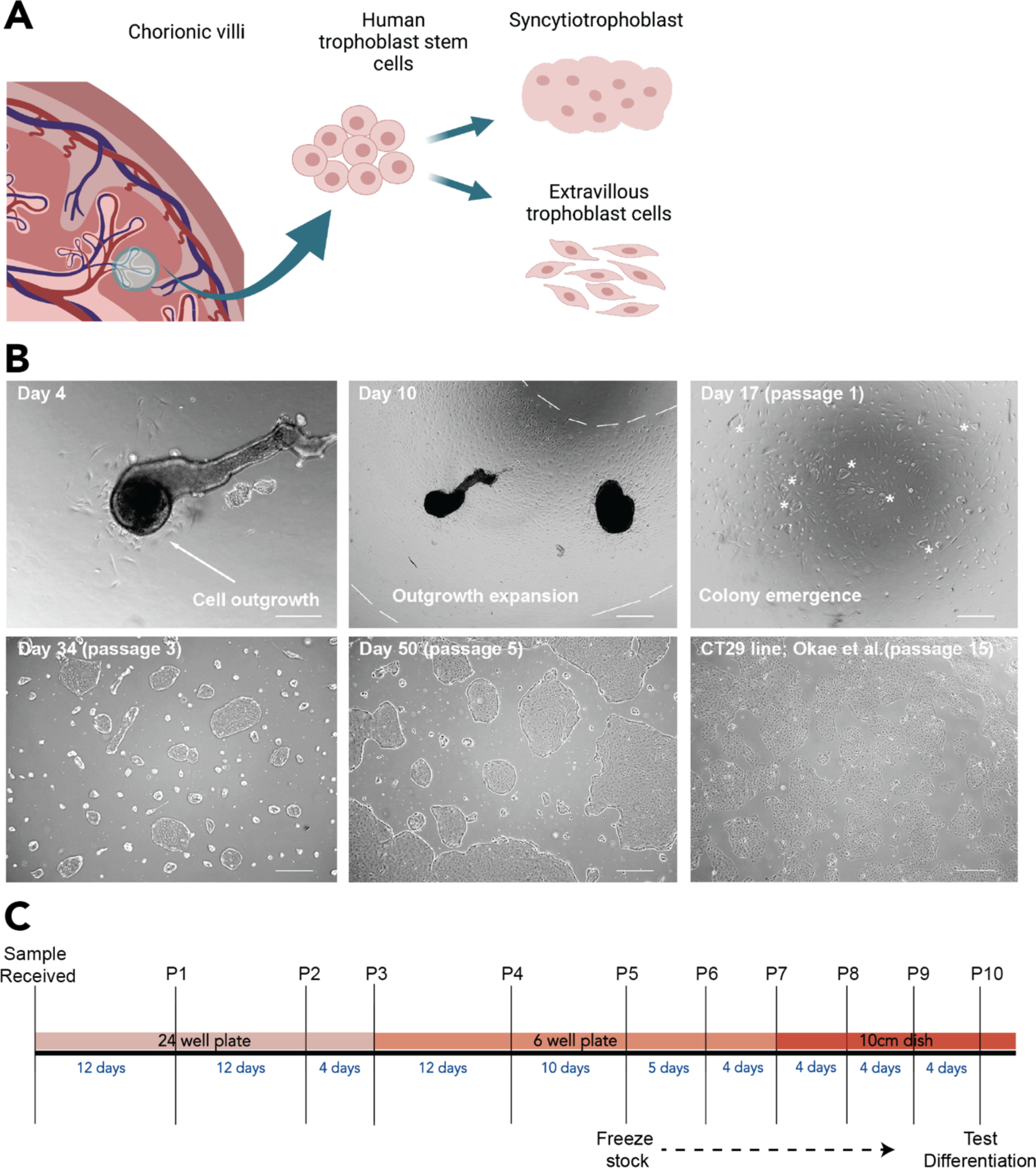
Deriving TS cells from chorionic villus tissue specimens. **A)** Simplified schematic depicting the process of obtaining chorionic villi tissue fragments, derivation of TS cells, and then subsequent differentiation into STB and EVT cell lineages. Created with BioRender.com. **B)** Chorionic villus tissue fragments attach and form cellular outgrowths within a few days of initial plating. Within one to two weeks the outgrowths expand and proliferate across the well. Two to three weeks after plating, the cells were passaged, and colonies emerged. Colony clusters were initially small but proliferated and grew rapidly. Significant heterogeneity is present initially, but subsequent passaging selects for a TS cell population that displays a similar morphology to the original TS cell lines (Okae et al., 2018), which possess the ability to differentiate into STB and EVT cell lineages. Scale bar represents 250 μm in first panel. All other scale bars represent 500 μm. **C)** An example timeline for TS cell line derivation and characterization.

**Fig. 2.**
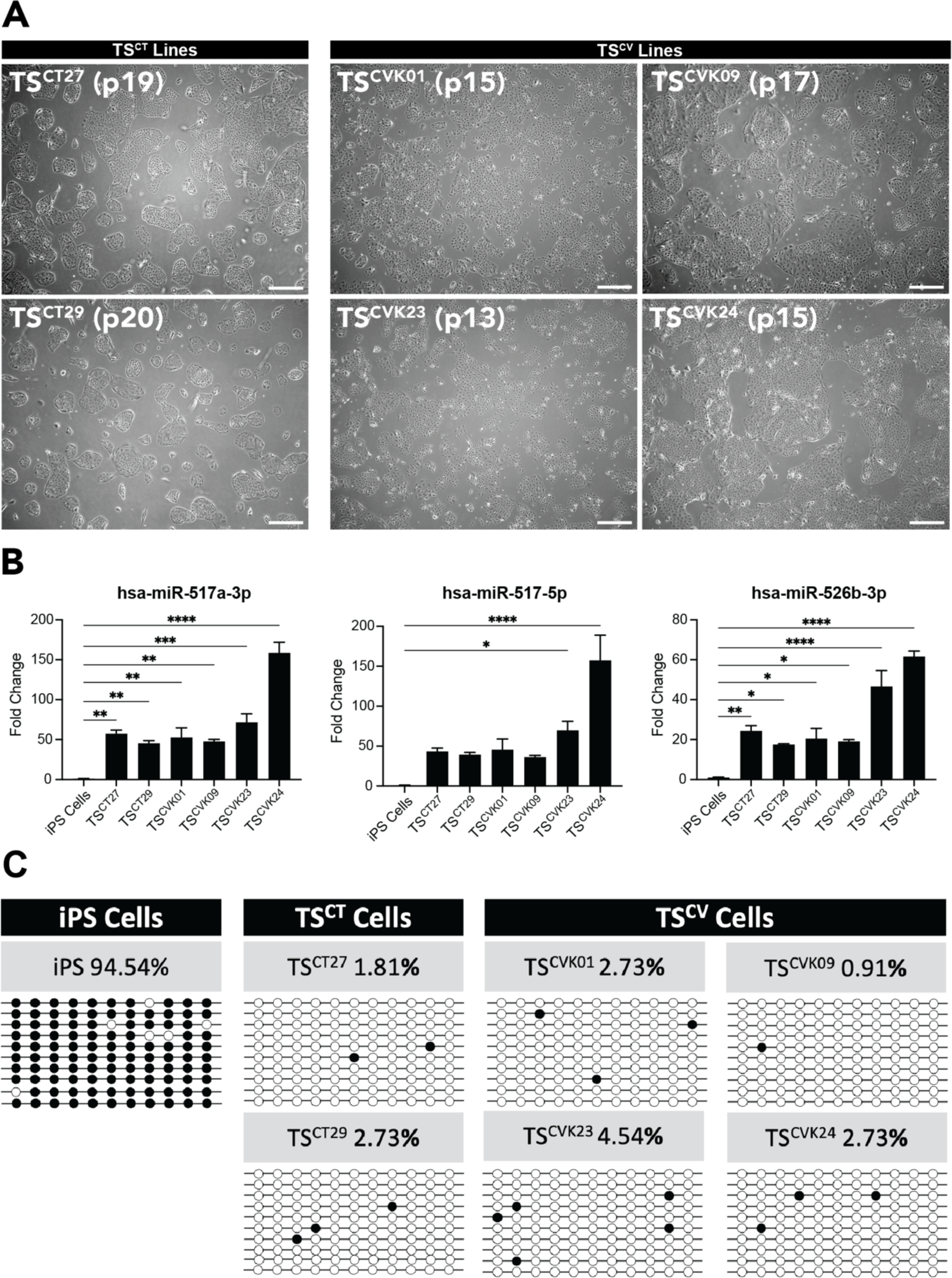
Characterization of TS^CV^ cells. **A)** Stem state phase contrast images of four chorionic villus-derived TS cell lines (TS^CVK01^, TS^CVK09^, TS^CVK23^, TS^CVK24^) alongside images of the reference cytotrophoblast-derived TS cell lines (TS^CT27^ and TS^CT29^) at different passage numbers (13-20). Scale bars represent 500 μm. **B)** Bar graphs depicting expression of three microRNAs (**miR**) from the C19MC cluster (hsa-miR-517a-3p, has-miR-517-5p, and hsa-miR-526b-3p) in TS^CT^ and TS^CV^ cell lines relative to induced pluripotent stem (**iPS**) cells, measured by RT-qPCR. Data were normalized to the control miRNA, hsa-miR-103a-3p (n=3 samples per group; *p<0.05, **p<0.01, ***p<0.001, ****p<0.0001). **C)** Plots representing DNA methylation levels in the ELF5 promoter at 11 sites in TS^CT^ and TS^CV^ cell lines compared to iPS cells. Methylated sites (black) and unmethylated sites (white) are shown for 10 replicates and the average percent methylation is listed.

The success rate of TS^CV^ line derivation was 42%, with ten TS^CV^ lines (6 XY; 4 XX) successfully derived from 24 unique patient tissue specimens (15 XY; 9 XX; **Table S1**). Success in cell line derivation may be impacted by the negative consequences of overnight shipping or the cellular contents of the tissue fragments but did not appear to be associated with the clinical karyotype of the CVS specimens. Maternal age range was 30-46 with a mean age of 37.3 years. Maternal and paternal ancestries were self-reported and varied (**Table S1**). Chorionic villus specimen collection ranged from 10-14 weeks gestation with an average gestation age of 12 weeks, 3 days (87 days). The earliest sample was collected at 10 weeks, 6 days gestation and the latest sample was collected at 14 weeks of gestation. The amount of starting tissue available for cell line derivation ranged from 5-20 mg with a mean of 8.5 mg of tissue. Clinical genetics performed on tissue specimens at the time of CVS indicated that 19 samples had normal karyotypes (13 samples 46, XY and 6 samples 46, XX). Clinical karyotyping of the remaining five samples reported genomic abnormalities including two samples with trisomy 18 (47, XX, +18 and 47, XY, +18), one sample with trisomy 21 (47, XX, +21), and two samples with chromosomal translocations (**Table S1**). The ten derived TS^CV^ cell lines were from eight samples with clinically normal karyotypes (TS^CVK01^, TS^CVK05^, TS^CVK09^, TS^CVK19^, TS^CVK21^, TS^CVK22^, TS^CVK23^, and TS^CVK24^), one with trisomy 21 (TS^CVK08^) and one with a translocation (TS^CVK07^). Additional phenotypic characterization was performed on a subset of TS^CV^ cell lines with 46, XX or 46, XY karyotypes.

### Characterization of TS^CV^ cells in the stem state

Karyotyping was repeated on TS^CV^ cells following line derivation and expansion. Cell line karyotypes were largely consistent with the clinical karyotyping (**Table S1**). Karyotypes of TS^CVK01^ and TS^CVK24^ lines were normal and consistent with the clinical results. TS^CVK09^ and TS^CVK23^ cell lines exhibited mosaicism and were not consistent with the clinical results. A subset of the cells karyotyped for TS^CVK09^ were 46, XY. The remaining cells analyzed displayed other genetic anomalies; however, each individual anomaly was restricted to 1-2 total cells (**Table S1**). A subset of TS^CVK23^ cells were identified as mosaic for trisomy 20 (47, XX, +20) following cell line derivation.

TS^CT27^ (XX) and TS^CT29^ (XY) served as reference standard human TS cell lines (Okae et al., 2018) used for comparative characterization of the TS^CV^ cell lines. TS^CV^ cells were maintained in a stem/proliferative state and propagated beyond the Hayflick limit of 50 cell divisions for non-stem cells, which is consistent with TS^CT^ cell proliferation (Okae et al., 2018). Established TS^CV^ cell lines were assessed for presence of mycoplasma by quantitative polymerase chain reaction (**qPCR**) and no evidence of contamination was detected. TS^CV^ cells in the stem state grew in discrete colonies and displayed a cobblestone morphology, consistent with the morphology of cytotrophoblast-derived cell lines, TS^CT27^ and TS^CT29^ ((Okae et al., 2018); **Fig. 2A**). TS^CV^ cells displayed additional characteristics consistent with their trophoblast cell identity (Lee et al., 2016), including expression of microRNAs from the Chromosome 19 microRNA cluster (**Fig. 2B; C19MC**; hsa-miR-517a-3p, has-miR-517-5p, and hsa-miR-526b-3p) and hypomethylation of the E74 Like ETS Transcription Factor 5 (***ELF5***) promoter relative to induced pluripotent stem (**iPS**) cells (**Fig. 2C**). Overall, TS^CV^ and TS^CT^ cells cultured in the stem state displayed similar proliferative, morphologic, microRNA expression, and methylation properties.

### Analysis of the differentiation capacity of TS^CV^ cells

Comparisons of TS^CV^ and TS^CT^ cell capacities for differentiation into STB and EVT cell lineages, were performed following cell line derivation (**Fig. 1C**). Assessments of cell differentiation were routinely performed following 10 passages. Differentiation was assessed at morphological and functional levels.

#### STB differentiation

The ability of TS^CV^ cells to differentiate into STB was assessed using the previously described three-dimensional STB (**ST3D**) protocol (Okae et al., 2018). STB differentiation elicited significant morphological changes, including the formation of suspended spheroid cell clusters (**Fig. 3A**). Complementary to the morphological changes observed, TS^CV^-derived STB displayed downregulation of stem state transcripts ***TEAD4***, ***LRP2***, and ***LIN28A*** (**Fig. 3B**) and upregulation of STB lineage-specific transcripts, including cytochrome P450 Family 11 Subfamily 1 (***CYP11A1***), chorionic gonadotropin beta 7 (***CGB7***), and syndecan 1 (***SDC1***; **Fig. 3C**). STB differentiated from TS^CV^ cells secreted chorionic gonadotropin (**CG**) at levels comparable to STB differentiated from TS^CT^ cells as measured by ELISA (**Fig. 3D**). Overall, TS^CV^-derived STB had similar cell morphology, expression patterns of signature STB transcripts, and CG production that is observed in TS^CT^-derived STB.

**Fig. 3.**
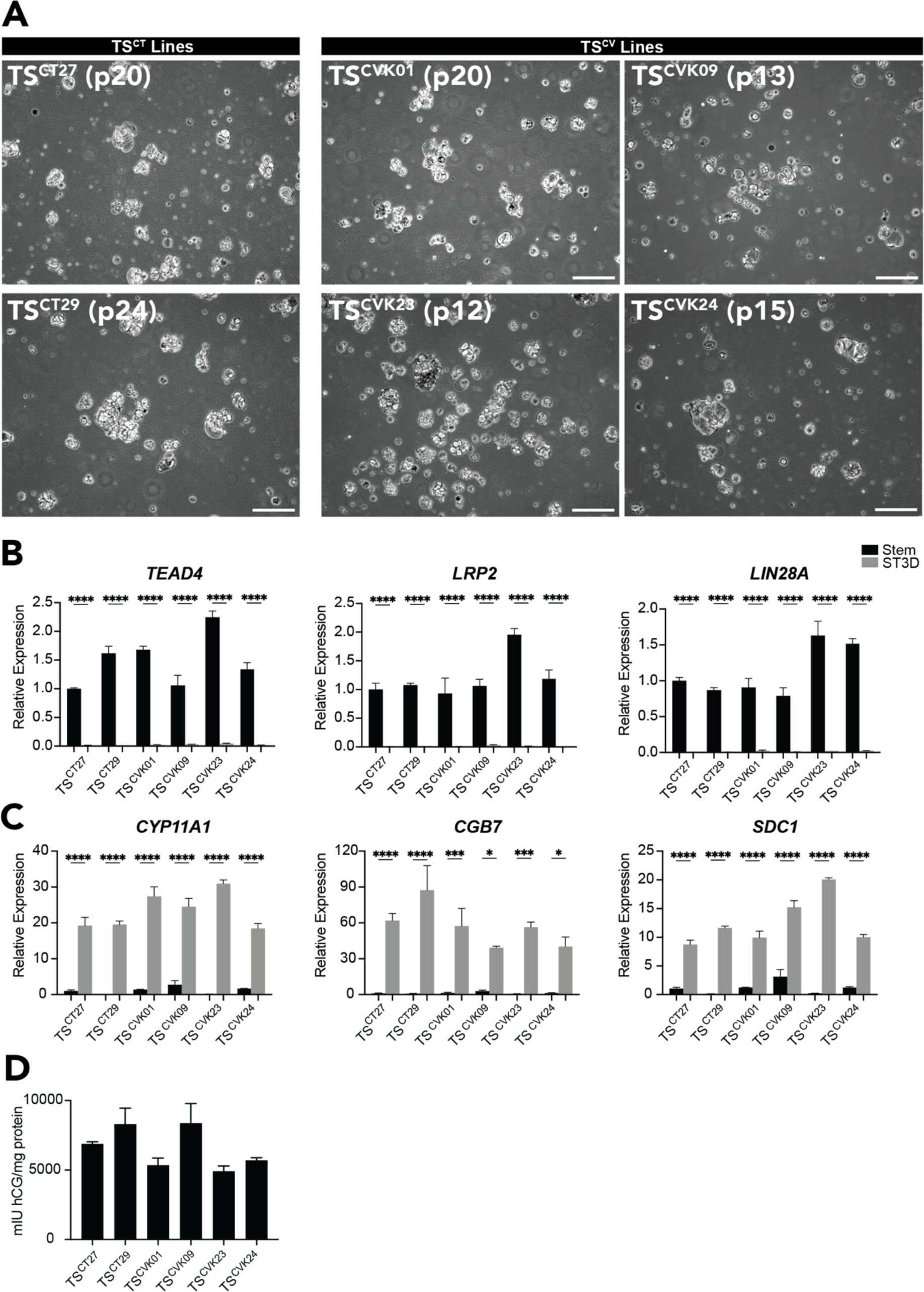
TS^CV^ cell line differentiation into STB. **A)** Representative phase contrast images of cytotrophoblast-derived TS^CT27^ and TS^CT29^ cells and four chorionic villus-derived TS cell lines possessing a normal karyotype, TS^CVK01^, TS^CVK09^, TS^CVK23^, and TS^CVK24^ cultured under STB differentiation conditions. Scale bars represent 250 μm. **B-C)** Stem cell-associated transcripts (**B;** *TEAD4*, *LRP2*, and *LIN28A*) and STB cell-associated transcripts (**C**; *CYP11A1, CGB7,* and *SDC1*) were quantified by RT-qPCR in stem (black) and STB differentiated (gray) TS^CT27^, TS^CT29^, TS^CVK01^, TS^CVK09^, TS^CVK23^, and TS^CVK24^ cells (n=3 samples per group; **p<0.01, ***p<0.001, ****p<0.0001). **D)** Chorionic gonadotropin (**CG**) protein levels (mlU/mg protein) were quantified by ELISA in cell culture supernatants collected from TS^CT^ and TS^CV^ cultured cells.

#### EVT cell differentiation

Canonical features of EVT cell differentiation observed in TS^CT^ cells were evident in TS^CV^ cell lines with normal karyotypes (TS^CVK01^, TS^CVK09^, TS^CVK23^, and TS^CVK24^), including elongated cell morphology (**Fig. 4A; Videos 1-3**) and expression of major histocompatibility complex, class I, G (**HLA-G**) protein (**Fig. 4B**). EVT cells displayed downregulation of stem state transcripts ***TEAD4***, ***LRP2***, and ***LIN28A*** (**Fig. 4C**). Characteristic EVT transcripts were upregulated, including *HLA-G*, matrix metallopeptidase 2 (***MMP2***), and C-C motif chemokine receptor 1 (***CCR1***; **Fig. 4D**). Overall, these TS^CV^ stem cell derived EVT cells were comparable to EVT cells derived from TS^CT^ cells.

**Fig. 4.**
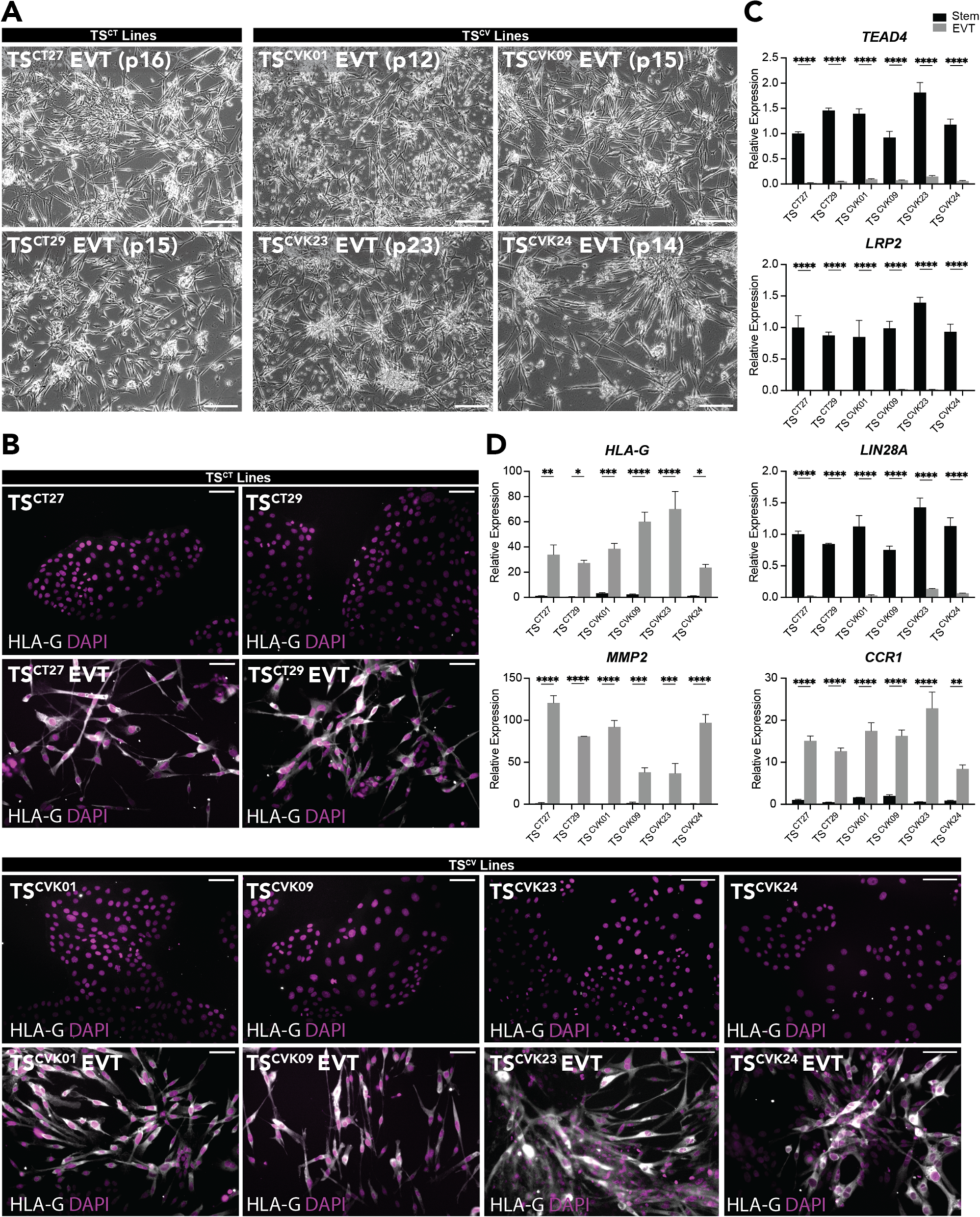
TS^CV^ cell line differentiation into EVT cells. **A)** Representative phase contrast images of cytotrophoblast-derived TS^CT27^ and TS^CT29^ cells and four chorionic villus-derived TS cell lines possessing a normal karyotype, TS^CVK01^, TS^CVK09^, TS^CVK23^, and TS^CVK24^ cells cultured under EVT cell differentiation conditions. Scale bars represent 250 μm **B)** Immunofluorescence detection of HLA-G (gray) by immunocytochemistry in TS^CT^ and TS^CV^ cells cultured in the stem state and on day 8 of EVT cell differentiation. DAPI (magenta) stains cell nuclei. Scale bars represent 100 μm. **C-D)** Stem cell-associated transcripts (**C;** *TEAD4*, *LRP2*, and *LIN28A*) and EVT cell-associated transcripts (**D**; *HLA-G*, *MMP2*, and *CCR1*) were quantified by RT-qPCR in stem (black) and EVT differentiated (gray) TS^CT27^, TS^CT29^, TS^CVK01^, TS^CVK09^, TS^CVK23^, and TS^CVK24^ cells (n=3 samples per group; *p<0.05, **p<0.01, ***p<0.001, ****p<0.0001).

### Transcriptomic analysis of the developmental potential of TS^CV^ cells

To obtain a broad comparative assessment of TS^CV^ and TS^CT^ in stem, STB, and EVT differentiated cell states, transcriptomes were captured using RNA-sequencing (**RNA-seq**). STB differentiation from the stem state resulted in broad changes in gene expression in TS^CT^ cells (TS^CT27^, TS^CT29^; **Fig. 5A; Table S2**) and TS^CV^ cells (TS^CVK01^, TS^CVK09^, TS^CVK23^, and TS^CVK24^; **Fig. 5B; Table S3**), including downregulation of stem markers *EPCAM, LIN28A, LRP2, PEG10,* and *TEAD4* and upregulation of STB markers *CGB2, CGB7, CYP11A1*, *CYP19A1,* and *SDC1*. STB differentiation-induced changes in gene expression were consistent between TS^CT^ (TS^CT27^ and TS^CT29^) and TS^CV^ (TS^CVK01^, TS^CVK09^, TS^CVK23^, and TS^CVK24^) cells (R=0.87, p<2.23-16; **Fig. 5C**).

**Fig. 5.**
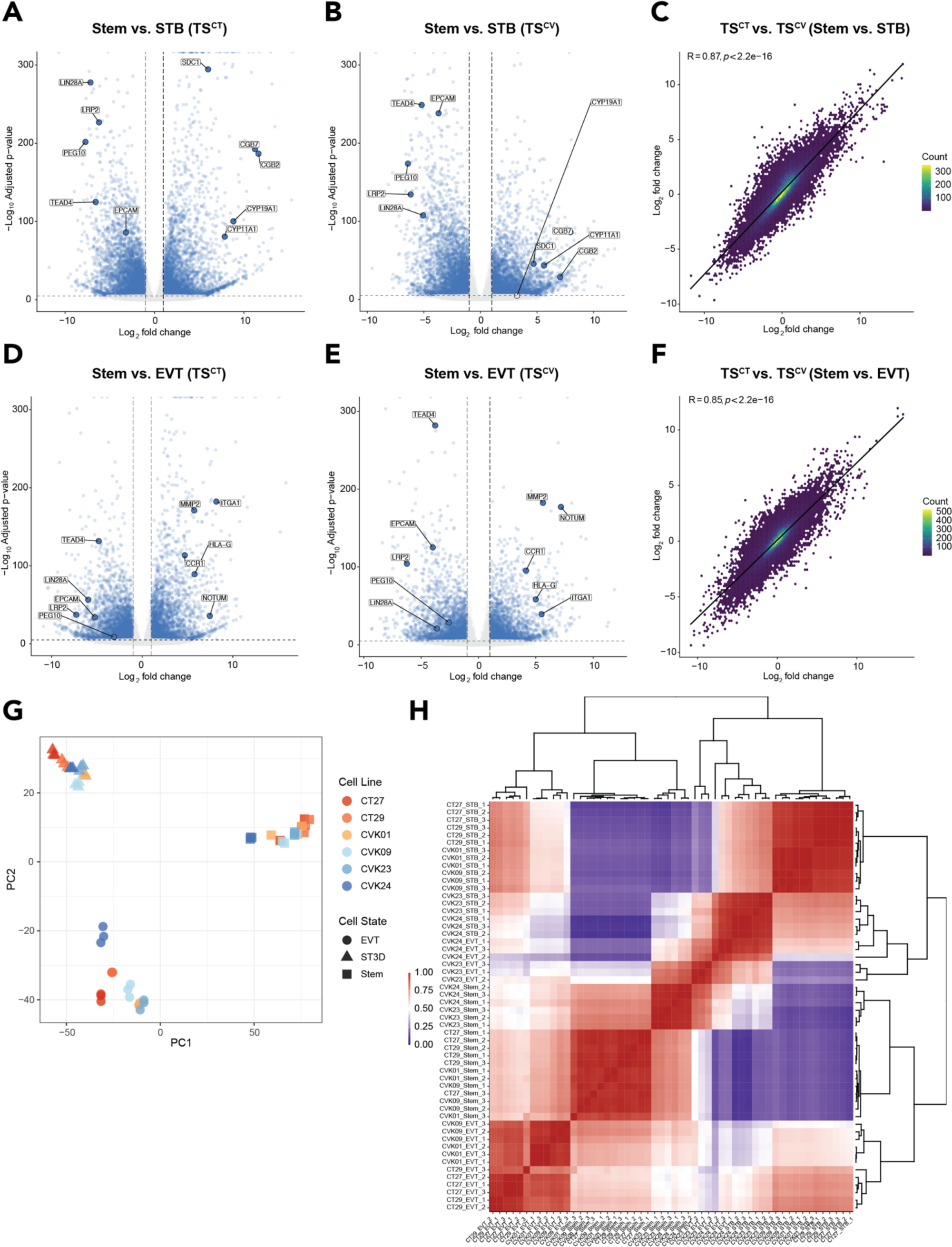
TS^CV^ and TS^CT^ cells cluster by cell state and share similar transcriptomes. **A-B)** Volcano plots depicting significantly up- and down-regulated genes based on transcripts measured by RNA-seq in stem versus STB state TS^CT^ (**A**) and TS^CV^ (**B**) cells. Gene transcript levels unchanged between stem and STB state cells are depicted in gray (n=3 per group; absolute Log_2_ fold change >1, adjusted p<0.05). **C)** Two-dimensional density plot comparing gene expression changes between stem and STB cell states in TS^CT^ (TS^CT27^ and TS^CT29^) cells versus TS^CV^ (TS^CVK01^, TS^CVK09^, TS^CVK23^, and TS^CVK24^) cells (Pearson correlation coefficient (R)=0.87, p<2.2e-16). **D-E)** Volcano plots depicting significantly up- and down-regulated genes based on transcripts measured by RNA-seq in stem versus EVT states of TS^CT^ (**E**) and TS^CV^ (**E**) cells. Gene transcript levels unchanged between stem and EVT state cells are depicted in gray (n=3 per group; absolute Log_2_ fold change >1, adjusted p<0.05). **F)** Two-dimensional density plot comparing gene expression changes between stem and EVT cell states in TS^CT^ (TS^CT27^ and TS^CT29^) cells versus TS^CV^ (TS^CVK01^, TS^CVK09^, TS^CVK23^, and TS^CVK24^) cells (Pearson correlation coefficient (R)=0.85, p<2.2e-16). **G)** Principal component analysis based on RNA-seq datasets generated from TS^CT^ and TS^CV^ cells cultured in the stem state or following differentiation into STB and EVT cell lineages. **H)** Heat map showing scaled normalized read counts representing gene expression profiles of stem state, STB, and EVT differentiated cells across TS^CT^ and TS^CV^ cell lines.

EVT cells successfully differentiated from the stem state exhibited broad gene expression changes in TS^CT^ (TS^CT27^, TS^CT29^; **Fig. 5D; Table S4**) and TS^CV^ cell lines with normal karyotypes (TS^CVK01^, TS^CVK09^, TS^CVK23^, and TS^CVK24^; **Fig. 5E; Table S5**). These changes included the downregulation of stem markers *EPCAM, LIN28A, LRP2, PEG10,* and *TEAD4* and upregulation of EVT cell markers *CCR1, HLA-G, ITGA1*, *MMP2,* and *NOTUM*. Gene expression changes induced by EVT cell differentiation were consistent between TS^CT^ (TS^CT27^ and TS^CT29^) and TS^CV^ (TS^CVK01^, TS^CVK09^, TS^CVK23^, and TS^CVK24^) cells (R=0.85, p<2.23-16; **Fig. 5F**).

Principal component analysis of TS^CT^ and TS^CV^ cell lines identified three primary cell-state specific clusters (**Fig. 5G**). TS^CV^ cells displayed consistent clustering in the stem state and following STB and EVT cell lineage differentiation (**Fig. 5G**). Results from correlation analyses performed to compare cell expression profiles are indicative of comparable transcriptomic changes across TS^CV^ and TS^CT^ cell lines (**Fig. 5H**). Overall, these results indicate that TS^CV^ cells are capable of self-renewal and effective differentiation into both STB and EVT cell lineages and can be considered bonafide TS cells.

## Discussion

Our understanding of placenta development and function has benefitted from the availability of in vitro model systems. In the human, these model systems have included primary cell and explant cultures, choriocarcinoma-derived cell lines, and immortalized cell lines (Ringler and Strauss, 1990, Shibata et al., 2020). Each in vitro approach has had merits but also limitations (Lee et al., 2016, Soares et al., 2018). Over two decades ago, Rossant and colleagues reported a procedure for culturing TS cells from the mouse (Tanaka et al., 1998). These cells could be maintained in a proliferative stem state or induced to differentiate. Furthermore TS cells could be reintroduced into blastocysts and shown to possess the capacity to contribute to mouse placentas (Tanaka et al., 1998). Mouse TS cells became an effective model system to elucidate gene regulatory networks controlling trophoblast cell differentiation and placental development (Hada et al., 2022, Hemberger et al., 2020, Latos and Hemberger, 2016, Lee et al., 2019). Efforts ensued to establish TS cells in other species with some success (Asanoma et al., 2011, Grigor’eva et al., 2009) but human TS cells represented an enigma (Kunath et al., 2014, Shibata et al., 2020). Culture protocols for sustaining mouse TS cells were ineffective in the human (Kunath et al., 2014). The discovery of culture conditions for propagating and differentiating human TS cells represented a major advancement (Okae et al., 2018). Utilizing these human TS cell culture tools, we have demonstrated the feasibility of capturing and expanding authentic TS cells from human chorionic villus specimens. Unique to these newly derived stem cells is the ability to obtain clinical outcomes that can be used to study placental development leading to healthy outcomes and disease states.

The initial human TS cell lines were derived from either blastocysts or first trimester pregnancy terminations (Okae et al., 2018). These human TS cell lines represent the benchmark for all TS cell lines subsequently derived. Chorionic villus biopsies are an alternative tissue source for deriving TS cells. They are retrieved during the first trimester of pregnancy as part of standard medical care (Adusumalli et al., 2007, Dong et al., 2003, McIntosh et al., 1993, Pisarska et al., 2016, Stranc et al., 1997, Wang et al., 1994, Williams et al., 1992, Williams et al., 1987). Thus, chorionic villus-derived TS cell lines can be connected to robust pregnancy outcome information. Human TS cell lines have also been derived from miscarriages (Saha et al., 2020), term human placenta tissue (Wang et al., 2022), and reprogrammed from pluripotent stem cells (Castel et al., 2020, Cinkornpumin et al., 2020, Dong et al., 2020, Guo et al., 2021, Io et al., 2021, Jang et al., 2022, Liu et al., 2020, Soncin et al., 2022, Viukov et al., 2022, Wei et al., 2021, Yanagida et al., 2021). These alternative TS cell models are potentially useful tools for investigating trophoblast cell development but each offer caveats for consideration. TS cells derived from trophoblast tissue obtained from miscarriages may best contribute to understanding trophoblast cell-related mechanisms linked to pregnancy failure and the impact of a failed pregnancy on TS cells. TS cells recovered from term placental tissue reflect the culmination of events transpiring throughout the duration of pregnancy, as epigenomic differences are observed in the placenta during gestation (Flowers et al., 2021, Gonzalez et al., 2021). Pluripotency is established through extensive genomic reprogramming (Hanna et al., 2010, Papp and Plath, 2013), which minimizes the impact of the epigenetic landscape established during pregnancy on the TS cell phenotype. It is reasonable to assume that genetic background and source of trophoblast tissue for TS derivation will influence TS cell behavior. Culturing TS cells under optimized conditions may normalize some features attributed to an adverse pregnancy and maternal environment, whereas in other cases the aberrant behavior may persist. Advantages of using chorionic villus-derived TS cells for investigating trophoblast cell-gene regulatory networks contributing to placental development are evident.

TS cell lines were successfully derived from chorionic villus biopsies possessing both normal and abnormal karyotypes. TS cells with a triploid karyotype have also been established from human blastocysts (Kong et al., 2022). Most recently, trophoblast organoids with abnormal karyotypes have been derived from chorionic villus biopsies (Schaffers et al., 2022). Chorionic villus-derived TS cell lines could be interrogated in the stem state and following differentiation into either STB or EVT cell lineages. The phenotypic and functional parameters evaluated revealed similarities between cytotrophoblast and chorionic villus-derived TS cells when cultured for stem state maintenance or following STB and EVT cell differentiation. Some differences in the capacity for EVT cell differentiation among chorionic villus-derived TS cells were noted. Variability in the capacity for human TS cell line differentiation into EVT cells has been previously reported (Cinkornpumin et al., 2020, Haider et al., 2022, Okae et al., 2018, Shannon et al., 2022). Numerous factors such as unreported clinical characteristics, undetected genomic differences, or inherent sample variability could be contributing to the differences observed. Thus, chorionic villus-derived TS cells represent a unique in vitro model to investigate functional variability in TS cells isolated from a temporally relevant tissue source. The true impact of the chromosomal abnormalities on TS cells and their differentiation into STB or EVT cells will require successful cultivation and characterization of multiple cell lines possessing the same abnormal karyotype.

Mosaicism is a characteristic feature of the human placenta (Coorens et al., 2021, Robinson and Del Gobbo, 2021, Yuen and Robinson, 2011). Trophoblast cells possess a tolerance for karyotypic abnormalities not evident in the embryo or fetus (Coorens et al., 2021, Shahbazi et al., 2020, Yuen and Robinson, 2011). Each cotyledon of the placenta exhibits elements of trophoblast cell clonality (Coorens et al., 2021). Placental mosaicism is manifested in genetic and functional differences among cotyledons within a human placenta (Coorens et al., 2021, Huang et al., 2009, Rubin et al., 1993, Wang et al., 1993). Among the TS cell lines derived from chorionic villus biopsies, some exhibited a karyotype consistent with the karyotype of chorionic villus tissue used for the clinical genetic analysis, whereas others differed. Chorionic villus biopsies contain a mixture of trophoblast and extraembryonic mesoderm (Aplin and Jones, 2021). Thus, differences in TS cell versus chorionic villus tissue could be attributed to confined placental mosaicism or alternatively, linked to an unappreciated consequence of culture conditions required to establish the TS cell lines.

In summary, the generation of chorionic villus biopsy-derived human TS cell lines expand the genetic diversity of existing TS cell lines available for basic research and importantly provides an opportunity to associate pregnancy outcomes with trophoblast cell biology. The research also offers insights into the significance of genetic anomalies and mosaicism in trophoblast cell development and introduces a novel precision medicine approach to the study of placentation.

## Materials and Methods

### Chorionic villus tissue collections, karyotypic analysis, and clinical phenotyping

Chorionic villus tissue was obtained by highly experienced perinatologists as part of standard medical care between 10-14 weeks of gestation for clinical genetic diagnosis at Cedars-Sinai Medical Center (Huang et al., 2009, Pisarska et al., 2016). Clinical cytogenetic analysis was performed on tissue specimens by direct and long-term culture and reviewed by a team of cytogeneticists (Huang et al., 2009). Residual trophoblast tissue fragments not required for clinical cytogenetic analysis were recovered, suspended in Complete TS Cell Medium [DMEM/F12 (11320033, Thermo Fisher, Waltham, MA), 100 μM 2-mercaptoethanol, 0.2% (vol/vol) fetal bovine serum (**FBS**), 50 μM penicillin, 50 U/mL streptomycin, 0.3% bovine serum albumin (**BSA**, BP9704100, Thermo Fisher), 1% Insulin-Transferrin-Selenium-Ethanolamine (**ITS-X**) solution (vol/vol, 51500056, Thermo Fisher)], 8.5 μM L-ascorbic acid (A8960, Sigma-Aldrich, St. Louis, MO), 50 ng/mL epidermal growth factor (**EGF**, E9644, Sigma-Aldrich), 2 μM CHIR99021 (04-0004, Reprocell, Beltsville, MD), 0.5 μM A83-01 (04-0014, Reprocell), 1 μM SB431542 (04-0010, Reprocell), 800 μM valproic acid (P4543, Sigma-Aldrich), and 5 μM Y27632 (04-0012-02, Reprocell)] (Okae et al., 2018), shipped overnight to the University of Kansas Medical Center, and used for TS cell derivation. Demographic data was collected from patients and included, parental ages, races and ethnicities, and ancestry (**Table S1**).

### Derivation of TS cells from chorionic villus tissue specimens

Chorionic villus biopsy tissue fragments were dissected and transferred in complete human TS cell medium. Briefly, individual villus fragments were minced and transferred to a 1.7mL tube, washed with PBS, and centrifuged at 500 x g for 3 min. Tissue pellets were resuspended in HBSS (with Ca^2+^ and Mg^2+^) supplemented with 1.25U/ml dispase II, 0.4mg/ml collagenase IV and 80 U/ml DNase I. Samples were then agitated for 15 min at 37°C. After incubation, tissue suspensions were centrifuged at 500 x g for 3 min. and washed with basal TS cell medium. Finally, cells and tissue suspensions were centrifuged at 500 x g for 3 min, resuspended in complete human TS cell medium and plated in 5 mg/mL Corning® mouse type IV collagen (35623, Discovery Labware Inc., Billerica, MA) coated dishes containing complete human TS cell medium. Cells and remaining tissue fragments attached within 2-5 days. Medium was replaced with fresh TS cell culture medium after initial attachment and every two days thereafter. Time to first passage was unique to each sample and determined by the extent of the outgrowth, but commonly occurred around 21 days post plating. Cells and attached tissue fragments were washed with PBS and detached with TrypLE Express (12604021, Thermo Fisher). Cell and tissue fragments were replated in human TS cell culture conditions in a 24 well plate format. Colonies emerged after the first passage. Cells were maintained in 24 well plate format for 3-5 passages and then expanded into 6 well plate format.

### TS cell culture

Following TS cell derivation, TS cells were cultured in dishes pre-coated with iMatrix511 (1:2000 dilution; NP892-01, Reprocell). TS cells were maintained in Modified Complete TS Cell Medium [DMEM/F12 (11320033, Thermo Fisher), 50 U/mL penicillin, 50 μg/mL streptomycin, 0.15% BSA (BP9704100, Thermo Fisher), 1% ITS-X solution (vol/vol; 51500056, Thermo Fisher)], 200 μM L-ascorbic acid (A8960, Sigma-Aldrich), 1% KnockOut Serum Replacement (**KSR**, 10828028, Thermo Fisher), 25 ng/mL EGF (E9644, Sigma-Aldrich), 2 μM CHIR99021 (04-0004, Reprocell), 5 μM A83-01 (04-0014, Reprocell), 800 μM valproic acid (P4543, Sigma-Aldrich), and 2.5 μM Y27632 (04-0012-02, Reprocell)] (Takahashi et al., 2019) medium was replaced every two days of culture. TS^CT27^ (XX) and TS^CT29^ (XY) (Okae et al., 2018) were used as reference lines.

### STB differentiation

To induce STB cell differentiation, TS cells were plated into 6 cm petri dishes at a density of 300,000 cells per dish and cultured in ST3D Medium [DMEM/F12 (11320033, Thermo Fisher), 50 U/mL penicillin, 50 μg/mL streptomycin, 0.15% BSA (BP9704100, Thermo Fisher), 1% ITS-X solution (vol/vol; 51500056, Thermo-Fisher)], 200 μM L-ascorbic acid (A8960, Sigma-Aldrich), 5% KSR (10828028, Thermo Fisher), 2.5 μM Y27632 (04-0012, Reprocell), 2 μM forskolin (F6886, Sigma-Aldrich), and 50 ng/mL of EGF (E9644, Sigma-Aldrich)](Okae et al., 2018). On day 3 of cell differentiation, 3 mL of fresh ST3D medium was added to the culture dishes. Cells were analyzed on day 6 of STB cell differentiation.

### EVT cell differentiation

EVT cell differentiation was induced by plating human TS cells onto 6-well plates pre-coated with 1 μg/mL of mouse type IV collagen at a density of 80,000 cells per well. Cells were cultured in EVT Differentiation Medium [DMEM/F12 (11320033, Thermo Fisher), 100 μm 2-mercaptoethanol, 50 U/mL penicillin, 50 μg/mL streptomycin, 0.3% bovine serum albumin (BP9704100, Thermo Fisher), 1% Insulin-Transferrin-Selenium-Ethanolamine solution (vol/vol; 51500056, Thermo Fisher)], 100 ng/mL of neuregulin 1 (**NRG1**, 5218SC, Cell Signaling, Danvers, MA), 7.5 μM A83-01 (04-0014, Reprocell), 2.5 μM Y27632 (04-0012, Reprocell), 4% KSR (10828028, Thermo Fisher), and 2% Matrigel^®^ (CB-40234, Thermo Fisher). On day 3 of EVT cell differentiation, the medium was replaced with EVT Differentiation Medium excluding NRG1 and with a reduced Matrigel^®^ concentration of 0.5%. On day 6 of EVT cell differentiation, the medium was replaced with EVT Differentiation Medium with a Matrigel^®^ concentration of 0.5% and excluding NRG1 and KSR. Cells were analyzed on day 8 of EVT cell differentiation.

### Cell line karyotyping

Chromosome analysis of TS^CV^ cells was performed using standard cytogenetic methods (Huang et al., 2009, Pisarska et al., 2016). GTG banded chromosomes were analyzed at 450-550 band levels. Cytogenetic and fluorescence in situ hybridization results were described according to the current International Standing Committee on Human Cytogenetic Nomenclature (ISCN, 2009).

### Immunocytochemical analysis

Cells were fixed with 4% paraformaldehyde (Sigma-Aldrich) for 15 min at room temperature. Fixed cells were incubated with primary antibody against HLA-G (ab52455, Abcam), followed by Alexa488-conjugated goat-anti-mouse immunoglobulin G (**IgG**; A32723, Thermo Fisher Scientific) secondary antibody and 4′,6-diamidino-2-phenylindole (**DAPI**; Molecular Probes, Eugene, OR). Fluorescence images were captured on a Nikon 80i upright microscope (Nikon) with a Photometrics CoolSNAP-ES monochrome camera (Roper Technologies, Inc., Sarasota, FL).

### iPS cell culture

Human iPS cells were propagated in tissue culture plates pre-coated with Matrigel^®^ (1:100 dilution; 356231, Corning Life Sciences, Tewksbury, MA). iPS cells were maintained in complete iPS Cell Medium [mTeSR1 Basal Medium + mTeSR1 5X Supplement (85850, STEMCELL Technologies, Inc., Vancouver, CA) and 10 μM Y27632 (04-0012-02, Reprocell)] and incubated at 37°C and 5% CO_2_. After the first day of culture, cells were cultured in complete iPS cell medium without Y27632. Medium was replaced every other day of culture. Cells were passaged or harvested at 80% confluency.

### miRNA isolation, cDNA preparation, and quantitative real-time PCR

Total RNA was isolated using mirVana kit (AM1560, Thermo Fisher), and RNA concentration was measured with the Qubit™ RNA BR Assay Kit (Thermo Fisher). cDNA synthesis was performed with TaqMan® Advanced miRNA cDNA Synthesis kit (A28007, Thermo Fisher). RT-qPCR was performed using TaqMan™Fast Advanced Master Mix (4444556, Thermo Fisher) and targeted miRNAs MIR517a-3p, MIR517-5p, MIR-526b-3p, and housekeeping miRNA MIR103a-3p (479485_mir, 478996_mir, and 478253_mir; TaqMan™ Advanced miRNA Assays, Thermo Fisher; **Table S6**). Relative expression of each transcript was calculated using ΔΔCT method and normalized to hsa-miR-103a-3p.

### Methylation analysis

Genomic DNA was isolated using the DNeasy Blood and Tissue Kit (69504, Qiagen, Germantown, MD), and 500 ng of DNA was bisulfite converted using the EZ DNA Methylation-Gold Kit (D5005, Zymo Research, Irvine, CA) according to instructions. Following bisulfite conversion, the ELF5 promoter region was amplified using a nested PCR approach with previously reported primers (Primer Set A: forward: 5’-GGAAATGATGGATATTGAATTTGA-3’; reverse: 5’-CAATAAAAATAAAAACACCTATAACC-3’ Primer Set B: forward: 5’-GAGGTTTTAATATTGGGTTTATAATG-3’; reverse: 5’-ATAAATAACACCTACAAACAAATCC-3’; **Table S7**; (Lee et al., 2016, Soncin et al., 2022). PCR was performed with a hot start DNA polymerase, ZymoTaq (E2001, Zymo Research). After the second PCR, Taq polymerase-amplified PCR products were gel-purified with QIAquick Gel Extraction Kit (28706X4, Qiagen), using manufacturer protocols. The purified DNA was inserted directly into a plasmid vector using TOPO® TA Cloning® Kits for Sequencing (450030, Thermo Fisher) according to manufacturer instructions. One microliter of purified PCR product was cloned into the plasmid vector (pCR™4-TOPO®) for 5 min at room temperature. Competent *E. coli* cells were transformed with the pCR4-TOPO construct, cultured, and minipreps were prepared using the QIAprep Spin Miniprep Kit (27106X4, Qiagen). Purified DNA was sequenced (GENEWIZ, South Plainfield, NJ).

### CG enzyme-linked immunosorbent assay (ELISA)

Conditioned medium was collected following six days of STB culture for each cell line. CG levels were measured using an ELISA kit (HC251F, Calbiotech), following the manufacturer’s protocol. The measurements were normalized to total cell protein content.

### RNA isolation and RT-qPCR

Total RNA was isolated using TRIzol^®^/chloroform precipitation (15596018, Thermo Fisher) as previously reported (Varberg et al., 2021). cDNA was synthesized from 1 μg of total RNA using the High-Capacity cDNA Reverse Transcription Kit (4368813, Thermo Fisher) and diluted 10 times with ultra-pure distilled water. qPCR was performed using PowerSYBR^®^ Green PCR Master Mix (4367659, Thermo Fisher) and primers (250 nM each). RT-qPCR primer sequences are presented in **Table S8**. Amplification and fluorescence detection were measured with a QuantStudio 5 Flex Real-Time PCR System (Thermo Fisher). An initial step (95°C, 10 min) preceded 40 cycles of a two-step PCR (92°C, 15 s; 60°C, 1 min) and was followed by a dissociation step (95°C, 15 s; 60°C, 15 s; 95°C 15 s). The comparative cycle threshold method was used for relative quantification of the amount of mRNA for each sample normalized to the housekeeping genes *B2M* or *POLR2A*.

### RNA library preparation and RNA-Seq

Stranded mRNA-sequencing was performed on the Illumina NovaSeq 6000 Sequencing System in the Genomics Core at the University of Kansas Medical Center. Quality control was completed with the RNA Screen Tape Assay kit (5067-5576, Agilent Technologies, Santa Clara, CA) on the Agilent TapeStation 4200. Total RNA (1 μg) was processed in the following steps: i) oligo dT bead capture of mRNA, ii) fragmentation, iii) reverse transcription, iv) cDNA end repair, v) Unique Dual Index (**UDI**) adaptor ligation, vi) strand selection, and vii) library amplification using the Universal Plus mRNA-Seq with NuQuant library preparation kit (0520-A01, Tecan Genomics, Männedorf, Switzerland). Library validation was performed with the D1000 Screen Tape Assay kit (5067-5582, Agilent Technologies) on the Agilent Tape Station 4200. Library concentrations were determined with the NuQuant module using a Qubit 4 Fluorometer (Thermo Fisher). Libraries were pooled based on equal molar amounts and the multiplexed pool was quantified, in triplicate, using the Roche Lightcycler96 with FastStart Essential DNA Green Master (06402712001, Roche, Indianapolis, IN) and KAPA Library Quant (Illumina, Inc., San Diego, CA) DNA Standards 1-6 (KK4903, KAPA Biosystems, Wilmington, MA). Using the qPCR results, the RNA-Seq library pool was adjusted to 2.125 nM for multiplexed sequencing. Pooled libraries were denatured with 0.2 N NaOH (0.04N final concentration), neutralized with 400 mM Tris-HCl, pH 8.0, and diluted to 425 pM. Onboard clonal clustering of the patterned flow cell was performed using the NovaSeq 6000 S1 Reagent Kit (200 cycle, 20012864, Illumina). A 2x101 cycle sequencing profile with dual index reads was completed using the following sequence profile: Read 1 – 101 cycles x Index Read 1 – 8 cycles x Index Read 2 – 8 cycles x Read 2 – 101 cycles. Sequence data were converted from .bcl to FASTQ file format using bcl2fastq software and de-multiplexed. Raw FASTQ files were trimmed using default parameters (-r 0.1 -d 0.03) in Skewer (Version 0.2.2) and reads shorter than 18 bp were discarded. Transcripts were quantified using Kallisto (Version 0.46.2). Differentially expressed genes (FDR of 0.05) were discovered using the Bioconductor package DESeq2 in R (Version 1.32.0).

### Live cell imaging

Cells were placed into an EVOS Onstage Incubator attached to an EVOS FL Automated Imaging System (Thermo Fisher). The live cell chamber was maintained at constant temperature (37°C), humidity, and 5% CO_2_. For stem culture, TS cells were maintained in stem state culture conditions described above and images were acquired 1-2 days after passage and immediately following culture medium change. EVT cell differentiation was induced as described above. On the fourth day of the EVT cell differentiation protocol, cells were placed into the live cell chamber. Phase contrast images were acquired every 10 min continuously from days 2-4 of stem cell growth or day 4-6 of EVT cell differentiation.

### Mycoplasma testing

Mycoplasma presence was assessed using the LookOut® Mycoplasma qPCR Detection Kit (MP0040A, Sigma-Aldrich). Kit protocols were followed as described.

## Data availability

All raw and processed sequencing data generated in this study have been submitted to the NCBI Gene Expression Omnibus (GEO; https://www.ncbi.nlm.nih.gov/geo/).

## Code availability

Only publicly available tools were used for data analysis and are described where relevant in the methods.

## Materials availability

Materials will be made available upon reasonable request to the investigators.

## Statistical analysis

Statistical analysis was completed with the GraphPad Prism 9 software. Welch’s *t* tests, Brown-Forsythe and Welch ANOVA tests were applied when appropriate. The figures depict the data represented as mean ± standard deviation with a statistical significance level of *p*<0.05.

## Study Approval

All human tissue specimens used for research purposes were collected following informed written consent, deidentified, and approved by institutional review boards at both Cedars-Sinai Medical Center and at the University of Kansas Medical Center.

## Data Availability

All data produced in the present study are available upon reasonable request to the authors.

## Acknowledgements

We thank Stacy Oxley and Brandi Miller for their administrative assistance and Clark Bloomer and Rosanne Skinner in the KUMC Genome Sequencing Facility for their work in library preparation and DNA sequencing. The research was supported by a National Institutes of Health (**NIH**) National Research Service Award postdoctoral fellowship (F32HD096809) and pathway to independence award (K99HD107262) to K.M.V., NIH grants (MDP: AI154535; MJS: HD020676, HD099638, HD105734), and the Sosland Foundation.

## Author contributions

K.M.V., A.M., M.D.P, and M.J.S. conceived and designed the research; A.N., J.W., and M.D.P. collected and obtained tissue specimens. J.G., I.S., H.O., and T.A. provided reagents, protocols, or equipment; K.M.V., A.M., B.M., and K.I., performed experiments and/or analyzed data; K.M.V, A.M., J.G, I.S., M.D.P, and M.J.S. interpreted results of experiments; K.M.V., A.M., J.M.V., and M.J.S. prepared figures and drafted manuscript; K.M.V., A.M., M.D.P, and M.J.S. edited and revised manuscript; All authors approved final version of manuscript.

## Diversity and inclusion statement

We are committed to promote diversity and inclusion in our research. Our team comprises individuals from varied backgrounds, and we seek to ensure equitable representation and recognition in our work and collaborations.

## Video Legends

**Video 1. Stem state and EVT-differentiated TS^CT27^ cell growth and motility.** Phase contrast images were acquired every 10 min for 53 h of TS^CT27^ cells cultured under stem state or EVT cell-differentiation conditions (EVT cell differentiation protocol days 4-6).

**Video 2. Stem state and EVT cell differentiated TS^CVK01^ cell growth and motility.** Phase contrast images were acquired every 10 min for 53 h of TS^CVK01^ cells cultured under stem state or EVT cell differentiation conditions (EVT cell differentiation protocol days 4-6).

**Video 3. Stem state and EVT cell differentiated TS^CVK09^ cell growth and motility.** Phase contrast images were acquired every 10 min for 53 h of TS^CVK09^ cells cultured under stem state or EVT cell differentiation conditions (EVT cell differentiation protocol days 4-6).

